# Evaluation of symptomatic patient saliva as a sample type for the Abbott ID NOW COVID-19 assay

**DOI:** 10.1101/2020.06.01.20119198

**Authors:** Jeffrey A. SoRelle, Lenin Mahimainathan, Clare McCormick-Baw, Dominick Cavuoti, Francesca Lee, Anjali Bararia, Abey Thomas, Ravi Sarode, Andrew E. Clark, Alagarraju Muthukumar

## Abstract

The severe acute respiratory syndrome coronavirus 2 (SARS-CoV-2) pandemic has presented significant challenges for laboratories including supply chain limitations with restricted access to reagents and sample collection materials (i.e. swabs, viral transport media (VTM)) for patients testing. Therefore, saliva has been evaluated as an alternative specimen for COVID-19 diagnosis. comparable performance between dry nasal swabs (NS) and nasopharyngeal swabs (NPS) collected in VTM has been observed with the ID NOW for SARS-CoV-2; the majority of false-negative results occur with higher cycle number (CN) or cycle threshold (Ct) values suggesting low viral load in these specimens. We performed clinical validation of saliva specimens on the ID NOW molecular platform to detect SARS-CoV-2. Saliva was compared to nasopharyngeal swabs tested on the ID NOW and the Cepheid molecular assay. We also performed stability studies of saliva samples over 5 days. A total of 96 saliva samples and 64 paired NPS were analyzed. We observed 78% (18/23) positive percent agreement (PPA) and 100% (44/44) negative percent agreement (NPA) between matched saliva and nasopharyngeal specimens performed on ID NOW. We found 83% (19/23) PPA and 100% NPA (25/25) between saliva run on the ID NOW and Cepheid assay. Six saliva specimens positive for SARS-CoV-2 were continuously positive for five days when stored at room temperature. Therefore, we propose further investigation of saliva as an alternative sample type for testing symptomatic patients with ID NOW as a promising method to address COVID-19 testing.

---

The severe acute respiratory syndrome coronavirus 2 (SARS-CoV-2) pandemic has presented significant challenges for laboratories including supply chain limitations with restricted access to reagents and sample collection materials (i.e. swabs, viral transport media (VTM)) for patients testing. Therefore, saliva has been evaluated as an alternative specimen for COVID-19 diagnosis. The collection of the saliva specimen does not require specialized materials, is non-invasive, and can be self-collected (1, 2).

Access to rapid and accurate testing is essential to limit the SARS-CoV-2 community spread and curtail COVID-19 resurgence. The Abbott ID NOW (ID NOW, Abbott Diagnostics, Sarborough, ME) is a point-of-care (POC) isothermal amplification-based platform that detects SARS-CoV-2 from patient specimens in approximately five minutes. ID NOW is currently used in pharmacies, hospitals, and various outpatient settings in all 50 states, and has a production capacity of 2 million tests per month by June 2020 (3).

Recently, concerns regarding ID NOW sensitivity compared to RT-PCR assays have been raised during clinical validations (4–7). The manufacturer has updated the specimen types indicated for use and the product information directing confirmatory testing with another molecular assay in cases of negative results with high clinical suspicion for COVID-19 (8). The comparable performance between dry nasal swabs (NS) and nasopharyngeal swabs (NPS) collected in VTM has been observed with the ID NOW for SARS-CoV-2 (4); the majority of false-negative results occur with higher cycle number (CN) or cycle threshold (Ct) values suggesting low viral load in these specimens (5, 7).

In the setting of limited specimen collection resources and an unknown optimal sample type for POC testing, we examined the performance of the ID NOW COVID-19 assay using symptomatic patient saliva. We tested a total of 96 patient saliva samples on the ID NOW. Sixty-seven specimens were paired collections with NPS in VTM. We first compared IDNOW saliva results with results from paired NPS specimens tested by either the Xpert® Xpress SARS-CoV-2 (Cepheid, Sunnyvale, CA) or Real-Time SARS-CoV-2 (Abbott Molecular, Des Plaines, IL) RT-PCR assays. All testing was performed per manufacturer’s instructions, with 200µL of saliva being used for ID NOW testing.

The limit of detection for the ID NOW was verified at 2000 copies/mL using synthetic RNA (SARS-CoV-2 standard, Exact Diagnostics, Ft. Worth, TX), consistent with previously published reports (7). We found a 78% (18/23) positive percent agreement (PPA) and 100% (43/43) negative percent agreement (NPA) for saliva tested by ID NOW compared with NPS in VTM (Table 1). False-negative (FN) saliva samples were associated with elevated NPS CN (Abbott, N2: 30.44) or Ct values (Cepheid, E: 33.0, 36.5, 40.6, 43.3; N2: 35.9, 36.1, 38.5, 39.0), consistent with previous reports for NPS (7).

**Table 1:** Saliva as sample type for ID NOW COVID-19 assay performance comparison against the Cepheid Real-Time PCR assay.

| Sample type | (n) | TP | FP | TN | FN | IN | PPA<br>% (95%CI) | NPA<br>% (95%CI) | PPV*<br>% | NPV*<br>% |
| --- | --- | --- | --- | --- | --- | --- | --- | --- | --- | --- |
| Saliva Vs NPS (RT-PCR) | 67 | 18 | 0 | 44 | 5 | 0 | 78 (56-93) | 100 (92-100) | 100 | 99 |
| Saliva Vs Saliva (Cepheid) | 49 | 19 | 0 | 25 | 4 | 1 | 83 (61-95) | 100 (86-100) | 100 | 99 |

\* PPV and NPV calculated based upon an institutional prevalence of 6.4% as determined through the analysis of all positive SARS-CoV-2 tests at this institution from 3/8-5/20/2020.
PPA: Positive percent agreement, NPA: Negative percent agreement, PPV: Positive predictive value, NPV: Negative predictive value, TP: True Positive, FP: False positive, TN: True negative, FN: False negative, IN: invalid

We recently validated the Cepheid assay for use with saliva (9) and compared the performance of ID NOW using 49 saliva specimens (Table 1). In this comparison, we observed 83% (19/23) PPA and 100% NPA (25/25). FN samples by ID NOW again exhibited elevated Ct values (E: 36.4, 36.5, 42.7, 43.3; N2: 36.1, 37.6, 39, 41.2). Chart review of all FN samples in this work revealed a majority (n = 4/7, 2 were common) were tested > 2 weeks after symptom onset, with most already receiving a diagnosis of COVID-19. Three of the FN patients had a previous diagnosis of COVID-19, another was diagnosed by ID NOW on an NPS specimen and retested while in the hospital, one was de-identified with no information and the last had no previous diagnosis but had an atypical presentation of weakness, and diarrhea for two weeks with subtle hazy opacities of the lung base indicative of minor respiratory disease.

Stability studies were undertaken to evaluate ID NOW performance as specimens aged(i.e. conditions simulating self-collection). NPS has exhibited reliable SARS-CoV-2 stability over a range of times and temperatures in a variety of storage media (10), but studies examining saliva are lacking. Six SARS-CoV-2-positive saliva specimens were evaluated by the ID NOW daily over 5-days post-collection stored at room temperature (22–25°C). All specimens were found to remain positive during serial testing with 200µL volumes over that time.

Resurgence in COVID-19 cases as social distancing and travel restrictions are relaxed necessitate expanded accessibility to POC devices. Saliva utilization for ID NOW testing supports self-collection and facilitates retesting of invalid or questionable results without the need for the recollection of a second NS or NPS due to specimen exhaustion. This could also limit the exposure of medical staff to potentially infectious patients. Therefore, we propose further investigation of saliva as an alternative sample type for testing symptomatic patients with ID NOW as a promising method to address COVID-19 testing.

## Data Availability

Data will be available for review upon request.

## Acknowledgments

The authors thank Julia Sweetnam MT(ASCP), Juancho Franco MT(ASCP), and Bryan Allen MT(ASCP) for excellent technical assistance.

